# Linezolid induced lactic acidosis in tuberculosis: a systematic review of published case reports, case series

**DOI:** 10.1101/2024.10.09.24315146

**Authors:** Rajiv Garg, Ravindra Kumar Garg, Pragati Garg, Anand Srivastava, Darshan Kumar Bajaj

**Author notes:** **Address for correspondence:** Dr. Rajiv Garg, Department of Respiratory Medicine, King George’s Medical University UP, Lucknow (India).

## Abstract

**Background:** Linezolid is widely used in the treatment of drug-resistant tuberculosis (TB), but it is associated with severe adverse effects, including lactic acidosis, which remains underreported in the literature and is fatal in many cases.

**Methods:** This systematic review followed PRISMA guidelines and was registered in PROSPERO (CRD42024552225). We conducted comprehensive searches across PubMed, Scopus, Embase, and Google Scholar, covering case reports and case series without language restrictions. Data were extracted on demographics, clinical presentation, Linezolid dosage, laboratory findings, treatment regimens, and outcomes. Lactic acidosis was defined as a serum pH <7.25 and lactate >4 mmol/L. Quality assessment followed the protocol of Murad MH et al. Data were synthesized narratively and statistically analyzed using Microsoft Excel.

**Results:** Seven case reports were identified. The average patient age was 49.33 years (range: 20–81 years), with a near-equal distribution between males and females. Most cases were reported from India (3/7). Respiratory symptoms predominated, with gastrointestinal and central nervous system involvement also noted. Linezolid dosing varied between 600 mg and 1200 mg daily. Arterial blood gas analysis showed significant acidosis, with pH values ranging from 6.09 to 7.32. Lactate levels were elevated (range: 10.2–20 mmol/L). Symptoms emerged within 7 days to over 1 month after starting Linezolid, except for one patient who developed symptoms immediately. Four patients died, and three survived after treatment modification.

**Conclusion:** Linezolid-induced lactic acidosis is a rare but serious complication in drug-resistant TB, emphasizing the need for close monitoring and individualized treatment.

## Introduction

Linezolid, a synthetic antibiotic from the oxazolidinone class, is crucial in managing drug-resistant tuberculosis (TB), particularly in multidrug-resistant (MDR) and extensively drug-resistant (XDR) cases. Originally developed to treat Gram-positive bacterial infections, Linezolid’s unique mechanism—blocking bacterial protein synthesis by inhibiting the initiation of translation—makes it effective against *Mycobacterium tuberculosis* strains resistant to both first- and second-line anti-TB drugs.^**1**^

As part of the National Tuberculosis Elimination Program (NTEP), Linezolid plays a significant role in both short and long MDR/XDR-TB treatment regimens, reflecting the increasing global concern over drug resistance. The World Health Organization (WHO) has classified Linezolid in Group A drugs for MDR-TB management, indicating its status as one of the most potent agents for resistant TB.^**2**^ However, its clinical use presents challenges due to a notable risk of toxicity, with up to 40% of patients experiencing side effects such as anemia, thrombocytopenia, and neuropathy. One of the more severe side effects is lactic acidosis.^**3**^

Lactic acidosis occurs due to the accumulation of lactate and protons in bodily fluids, typically following elevated blood lactate levels, termed hyperlactatemia. It is essential to differentiate between hyperlactatemia and lactic acidosis by considering factors like arterial blood gas values, electrolyte levels, and patient medical history. Lactic acid (LA) exists as two enantiomers, L-LA and D-LA. L-lactic acidosis (L-LA) is the more common form of metabolic acidosis, especially in critical care. D-lactic acidosis is rarer, occurring through a different metabolic pathway and typically produced by human microbiota. The definition of lactic acidosis varies, but it is generally regarded as a condition where L-lactate concentrations exceed 4 mmol/L in plasma, plasma bicarbonate is less than 20 mmol/L, and blood pH drops below 7.35. High concentrations of L-lactic acid can lead to severe acidemia, potentially causing cardiovascular collapse. Cellular dysfunction in these cases is thought to result from hydrogen ions binding to cellular proteins.^**4**^

In clinical settings, the reported incidence of lactic acidosis in patients treated with Linezolid ranges from 6.8% to 35.7%,varying based on factors like treatment duration, dosing, and patient-specific factors such as age and liver or kidney dysfunction. ^**5-6**^ The literature on Linezolid-associated lactic acidosis in drug-resistant TB patients is sparse, limited mostly to case reports and short communications. Given the rarity and seriousness of this condition, this systematic review was undertaken to evaluate the pattern and spectrum of Linezolid-induced lactic acidosis in patients with drug-resistant tuberculosis, based on published case reports and case series.

## Methodology

We conducted a systematic review of case reports and series that discuss Linezolid-induced lactic acidosis in cases of drug-resistant tuberculosis (TB), following the Preferred Reporting Items for Systematic Reviews and Meta-Analyses (PRISMA) guidelines. Our review protocol has been registered in PROSPERO, reference number [CRD42024552225].^**7**^

### Search Strategy

We performed a comprehensive search across multiple databases, including PubMed, Scopus, Embase, and Google Scholar. For Google Scholar, we reviewed the first 50 pages of search results. No language restrictions were applied, and for non-English articles, we used Google Translate to translate them into English. The search strategy employed was:

#### For PubMed

“Linezolid”[Mesh] OR “Linezolid” AND “Lactic Acidosis”[Mesh] OR “Lactic Acidosis” OR “Acidosis, Lactic” OR “Lactic Acid” OR “Lactate” AND “Case Reports” OR “case report” OR “case series” OR “cohort studies” OR “cohort study” OR “observational study” OR “retrospective study” OR “prospective study.”

#### For Embase

(‘linezolid’/exp OR ‘linezolid’) AND (‘lactic acidosis’/exp OR ‘lactic acidosis’ OR ‘acidosis, lactic’ OR ‘lactic acid’ OR ‘lactate’) AND (‘case report’/exp OR ‘case report’ OR ‘case series’ OR ‘cohort study’ OR ‘cohort studies’ OR ‘observational study’ OR ‘retrospective study’ OR ‘prospective study’). The last search was conducted on July 7, 2024.

### Definition

Lactic acidosis was defined as a serum pH <7.25 and serum lactate levels >4 mmol/L in patients with drug-resistant TB undergoing Linezolid treatment. The treatment duration was calculated from the initiation of the Linezolid-containing regimen.^**8**^

### Study Selection and Data Extraction

The review process was carried out in two phases. In the first phase, titles and abstracts were independently reviewed by two reviewers (RK and RG). Full-text articles were subsequently assessed based on predefined inclusion criteria. A third reviewer (AS) mediated any discrepancies. Quality assessment of the included studies was conducted by two additional reviewers (PG and SB), with a third opinion (DB) sought if needed. Records lacking complete information were excluded. Rayyan, a free web and mobile app for systematic reviews, was used for managing records and detecting duplicates, with oversight by two reviewers (RG and PG) and final arbitration by a third (RKG) when needed. The selection process was depicted using a PRISMA flowchart generated through Rayyan a web and mobile app for systematic reviews.^**9**^

Key information from selected case reports and series was extracted, including author details, publication year, patient demographics, clinical presentations, arterial blood gas results, type of TB, history of TB treatment, co-morbidities, details of the Linezolid-containing regimen, treatment duration, and outcomes related to Linezolid-induced lactic acidosis. This data collection was conducted by three reviewers (RG, AS, and RKG), with a fourth reviewer (DB) resolving any conflicts. Data was summarized in a series of tables, with statistical analyses performed using Microsoft Excel.

### Quality Assessment

Each case was evaluated using four primary criteria: selection, validation, causality, and reporting, in accordance with the protocol set by Murad MH et al.^**10**^ Case reports were classified as “good quality” if all criteria were met, “fair quality” if three criteria were met, and “poor quality” if only one or two criteria were met, following Della Gatta et al.’s framework.^**11**^

### Data Synthesis

Data were synthesized narratively and qualitatively. Categorical data were expressed as frequencies and percentages, while continuous variables were summarized using means, medians, and ranges. The final systematic review presented this data in tables, providing a comprehensive synthesis of the patterns and outcomes of Linezolid-induced lactic acidosis in drug-resistant TB patients.

### Results

Our search identified seven reports on patients with tuberculosis who developed linezolid-induced lactic acidosis (LILA) (Table 1 and Table-2).^**12-18**^ The systematic review process is outlined in the PRISMA flow diagram (Figure 1). Five of the cases were of good quality, and two were fair, as detailed in Supplementary Item 1. Data from all seven cases were included in the analysis (see Supplementary Item 2 for the PRISMA checklist).

**Figure 1.**
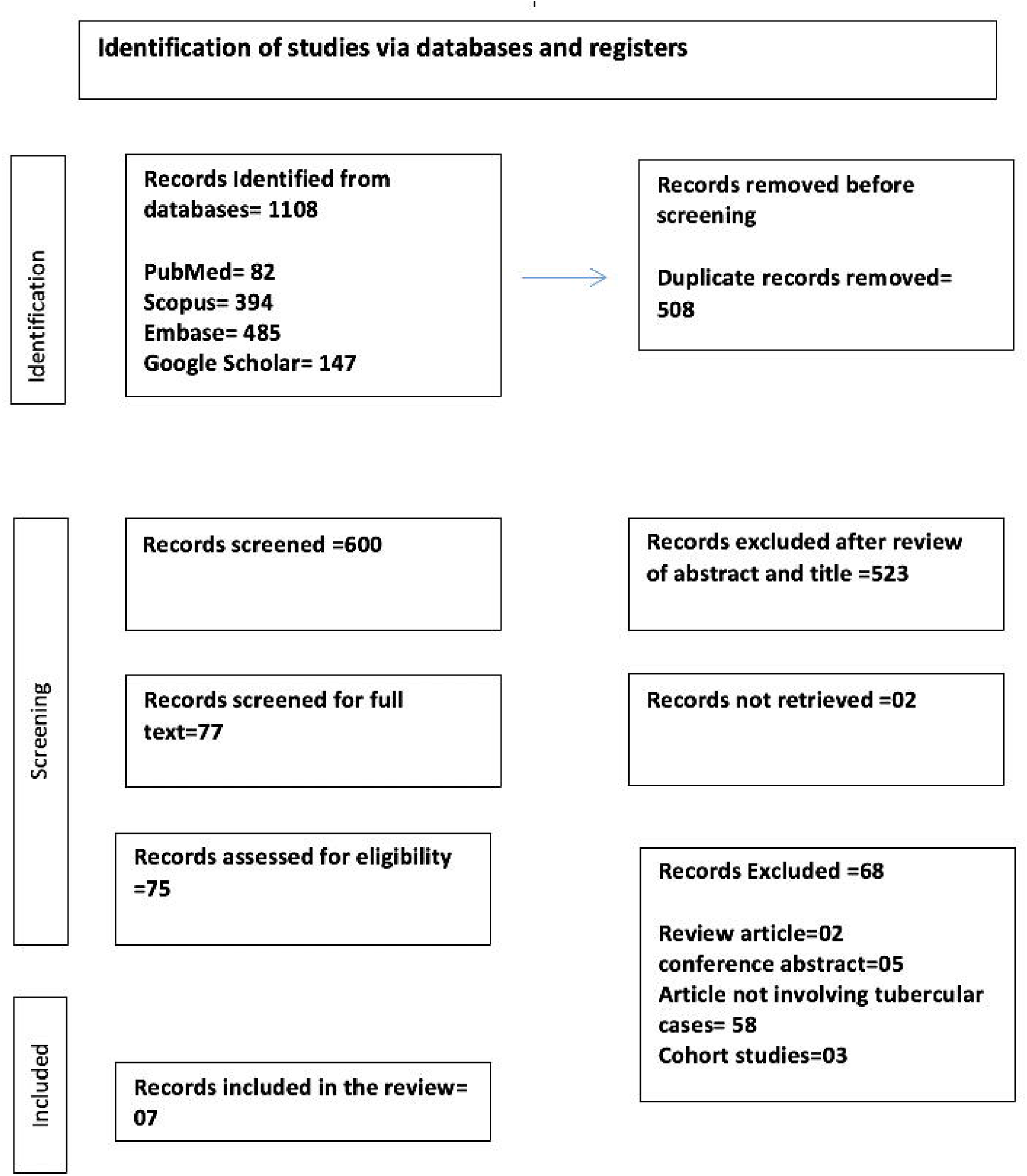
Flow diagram of the study

The patients’ average age was 49.33 years (median 47), with a range from 20 to 81 years. Three were male, and four were female. Geographically, cases were reported from India (3), Europe (2), South Africa (1), and China (1).

### Clinical Presentation

Respiratory symptoms were common (5/7 cases), followed by gastrointestinal symptoms (4/7). Two cases had central nervous system involvement. Pulmonary TB was seen in four patients, with one case each of disseminated TB, CNS TB, and vertebral TB with a psoas abscess.

### Treatment Regimens

Three patients were on 600 mg of Linezolid daily, three on 1200 mg, and one lacked dosage information. Treatment regimens included Bedaquiline, Levofloxacin, Cycloserine, and Clofazimine, with two patients having a prior history of anti-TB therapy.

### Laboratory Findings

Arterial blood gas analysis showed pH values ranging from 6.09 to 7.32 (mean 6.95, median 6.98). Lactate levels ranged from 10.2 to 20 mmol/L, with an average of 15.99 mmol/L.

### Onset of Symptoms

Symptom onset varied, with one case within a week, two between one week and a month, and four after one month.

### Outcomes

Linezolid was discontinued in three cases due to adverse effects. Two patients required ICU admission, and two underwent hemodialysis. Four patients died, while three recovered.

### Co-morbidities

Two patients had significant co-morbidities (HIV and kidney disease), while five had none documented.

### Drug Resistance

Five patients had multidrug-resistant TB (MDR-TB), while two cases (disseminated TB and TB meningitis) showed no resistance.

## Discussion

The findings from this review highlight the complexities involved in treating drug-resistant tuberculosis (TB), particularly with the use of Linezolid, a key second-line drug in multidrug-resistant (MDR) and extensively drug-resistant (XDR) TB management. While Linezolid is a powerful option against resistant TB strains, it carries a significant risk of severe adverse effects, including lactic acidosis, which poses a major challenge in clinical practice.

Lactic acidosis, characterized by low pH and high lactate levels, is a life-threatening condition that can lead to organ dysfunction and, as observed in this review, a high mortality rate. The wide range of symptoms observed, including respiratory, gastrointestinal, and central nervous system involvement, suggests that Linezolid-induced lactic acidosis (LILA) can present with diverse clinical features, further complicating diagnosis and management. Early detection of lactic acidosis is crucial, but the variability in the onset of symptoms after Linezolid initiation makes this difficult, as symptoms in some patients developed within the first week, while others appeared after more than a month of treatment.^**19**^

Another challenge noted is the need for individualized dosing and careful selection of concomitant drugs, as patients receiving both 600 mg and 1200 mg of Linezolid experienced severe side effects. This suggests that while dose reduction may mitigate some toxicities, it does not eliminate the risk of life-threatening complications. The need for ICU care and interventions like hemodialysis in some patients further highlights the seriousness of these cases.^**5**^

The presence of co-morbidities such as HIV and chronic kidney disease in some patients further complicates treatment and increases the risk of adverse outcomes. The fact that four out of seven patients in this cohort died underscores the importance of continuous monitoring, timely management of side effects, and the potential need to discontinue Linezolid when severe toxicity arises. ^**20**^

The study is limited by the small number of reported cases, lack of randomized control trials, and potential variability in reporting quality across case reports. Additionally, incomplete data on some patient outcomes and co-morbidities may affect the generalizability of the findings.

In conclusion, this review emphasizes the need for vigilant monitoring and personalized treatment plans when using Linezolid in drug-resistant tuberculosis cases. The high mortality rate and significant adverse effects observed in this small cohort underline the importance of balancing efficacy with safety and considering alternative treatment strategies when necessary.

## Data Availability

All data lies in the manuscript or supplementary material

## Declarations

### Ethical Statement

Since no human or animal subjects were involved, ethical clearance was not required.

### Data Availability Declaration

All data is available within the manuscript and supplementary files.

