## Supplementary material for "Linezolid induced lactic acidosis in tuberculosis: a systematic review of published case reports, case series": 1

| **Author(s)** | **Table tool used for the evaluation of the methodological quality of case reports and case series** | | |
| --- | --- | --- | --- |
| P. Scotton et al.  (ITALY)  P. Scotton,et al.  Infection 2008; 36: 387–388  DOI 10.1007/s15010-008-7329-3 | 1. Does the patient represent the whole experience of the investigator (center) or is the selection method unclear to the extent that other patients with similar  presentation may not have been reported? | First case where linezolid use for tuberculosis reported to have develop lactic acidosis. Case was of disseminated tuberculosis case with involvement of the meninges, mediastinum lymphnodes and bilateral pleural effusion. Pus was aspirated from paravertebral abscess by CT guidance and sent for bacterial and mycobacterial isolation and was confirmed to have M.Tb complex. | GOOD Quality |
|  | 2. Was the exposure adequately ascertained? | YES, The exposure to linezolid was clearly established. The patient received intravenous linezolid 600 mg twice daily as part of their treatment regimen for tuberculous spondylodiscitis. The dosage and administration route are clearly documented. |  |
|  | 3. Was the outcome adequately ascertained? | YES, The outcome, lactic acidosis, was adequately ascertained through objective measures. The diagnosis was made based on an arterial blood gas test revealing acidosis (pH 7.24), low bicarbonate (14.3 mmol/L), a high anion gap (24 mEq/L), and a high lactate level (18.6 mmol/L). These findings strongly support the presence of lactic acidosis. |  |
|  | 4. Were other alternative causes that may explain the observation ruled out? | No, The study acknowledges the potential for alternative explanations but doesn't definitively rule them out.  The authors acknowledge that isoniazid, another drug in the patient's antituberculous regimen, can induce lactic acidosis, particularly in overdose cases. However, they don't provide evidence ruling out an isoniazid overdose in this specific case.  The authors suggest that the severity of the tuberculous spondylodiscitis itself could contribute to lactic acidosis. However, they do not provide further details or evidence to support this claim. They don't explain how disease severity might lead to it. The patient's medical history, apart from the tuberculous spondylodiscitis, is not described. |  |
|  | 5. Was there a challenge and/or rechallenge phenomenon? | Yes, the introduction of linezolid to the patient's treatment regimen can be considered a challenge. The patient developed lactic acidosis after 12 days of receiving linezolid.  Rechallenge: No, Linezolid was not re-introduced. |  |
|  | 6. Was there a dose-response effect? | No, The study does not provide information about different linezolid doses or explore whether different dosages might be associated with varying risks of lactic acidosis. |  |
|  | 7. Was follow-up long enough for outcomes to occur? | Yes, In this specific case study, a follow-up of three weeks after the resolution of lactic acidosis and the reintroduction of isoniazid was deemed sufficient to demonstrate the patient's recovery and the absence of recurrent lactic acidosis. |  |
|  | 8. Is the case described with sufficient details to allow other investigators to replicate the research or to allow practitioners to make inferences related to their own practice? | Replication: does not offer enough detail for direct replication.  Inference: Yes, case study offers valuable insights for practitioners, in terms of awareness about the possibility of linezolid-associated lactic acidosis, even in cases of relatively short-term use, as was seen in this patient who developed the condition after 12 days of treatment.  It underscores the importance of monitoring patients on linezolid for signs and symptoms of lactic acidosis, particularly those with risk factors such as concurrent isoniazid use or severe illness.  Early detection: The authors suggest prompt checking of arterial blood gas and lactate levels in patients receiving linezolid who experience nausea and vomiting. This highlights the importance of early detection and intervention. |  |
| Letter to editor  David Boutoille et. al.  (FRANCE)  European Journal of Internal Medicine 20 (2009) e134–e135 | 1. Does the patient represent the whole experience of the investigator (center) or is the selection method unclear to the extent that other patients with similar  presentation may not have been reported? | The case study focuses on a 48-year-old man who developed lactic acidosis after prolonged linezolid exposure for multi-drug resistant tuberculosis (MDR-TB) treatment | GOOD Quality |
|  | 2. Was the exposure adequately ascertained? | Yes, the exposure to linezolid was clearly established. The patient received linezolid (600 mg twice daily) as part of a multi-drug regimen to treat MDR-TB. |  |
|  | 3. Was the outcome adequately ascertained? | Yes, the outcome, lactic acidosis, was confirmed through objective laboratory results. The patient presented with dyspnea and abdominal pain. An arterial blood gas test showed acidosis (pH 7.32), and his serum bicarbonate was low (14 mmol/L). The patient's arterial lactate was significantly elevated at 11.6 mmol/L, confirming the diagnosis of lactic acidosis., |  |
|  | 4. Were other alternative causes that may explain the observation ruled out? | Yes, The case study mentions that other common causes of lactic acidosis were ruled out. Use of alcohol, Metformin, liver failure, heart failure, sepsis all contributory causes were ruled out. |  |
|  | 5. Was there a challenge and/or rechallenge  phenomenon? | Challenge- No  Rechallenge-No deliberate rechallenge |  |
|  | 6. Was there a dose-response effect? | The case study focuses on a single patient receiving a standard dose of linezolid (600 mg twice daily). Therefore, it doesn't provide insights into the relationship between different linezolid doses and the likelihood or severity of lactic acidosis. |  |
|  | 7. Was follow-up long enough for outcomes to occur? | While the study mentions the patient was followed as an outpatient with monthly consultations, hemograms, and liver function tests, the follow-up duration relevant to the lactic acidosis event is limited. The patient died the day after presenting with symptoms and discontinuing linezolid. The study does not mention if the lactic acidosis was resolving at the time of his death. |  |
|  | 8. Is the case described with sufficient details to allow other investigators to replicate the research or to allow practitioners to make inferences related to their own practice? | It lacks the necessary details for direct replication.  It offers valuable insight for practitioners which highlights the potential for lactic acidosis as a complication of prolonged linezolid use, even in patients who show initial improvement and negative cultures.  It emphasizes the importance of regular monitoring for signs and symptoms of lactic acidosis in patients receiving linezolid, especially for extended durations. |  |
| Naiju Zhang  (CHINA)  Saudi Pharmaceutical Journal 30 (2022) 108–111 https://doi.org/10.1016/j.jsps.2021.12.021 | 1. Does the patient represent the whole experience of the investigator (center) or is the selection method unclear to the extent that other patients with similar  presentation may not have been reported? | The case report focuses on a 64-year-old Chinese woman who developed lactic acidosis after receiving linezolid for 28 days as part of her treatment for tuberculous meningitis | Good quality |
|  | 2. Was the exposure adequately ascertained? | Yes, the patient's exposure to linezolid was thoroughly documented. She received 600 mg of linezolid intravenously every 12 hours for 28 days before the onset of severe lactic acidosis. |  |
|  | 3. Was the outcome adequately ascertained? | Yes, the outcome, severe lactic acidosis, was confirmed with objective laboratory data. The patient's blood gas analysis revealed a pH of 6.944 (low), lactate of 16.5 mmol/L (high), and base excess of -27.3 mmol/L (low). These findings, coupled with her clinical symptoms of weakness, inability to walk, lethargy, decreased consciousness, drooping left eyelid, labored breathing, and dyspnea, led to the diagnosis of lactic acidosis, acute renal injury, and hyperkalemia. |  |
|  | 4. Were other alternative causes that may explain the observation ruled out? | The case report mentions that the patient was receiving a combination of antituberculosis drugs that could potentially cause lactic acidosis. However, the report doesn't offer a detailed account of how alternative causes were ruled out. The causal relationship was labelled as “Probable” in this case. |  |
|  | 5. Was there a challenge and/or rechallenge  phenomenon? | While not a traditional rechallenge, the reintroduction of antituberculosis drugs (excluding linezolid) after the patient's recovery and transfer to the infectious disease department did not result in a recurrence of lactic acidosis. This finding suggests that the lactic acidosis was primarily associated with linezolid and not the other antituberculosis drugs. |  |
|  | 6. Was there a dose-response effect? | The case report solely focuses on a single patient receiving a fixed dose of linezolid (600 mg every 12 hours). It doesn't provide insights into the potential relationship between varying linezolid doses and the likelihood or severity of lactic acidosis. |  |
|  | 7. Was follow-up long enough for outcomes to occur? | The case report primarily concentrates on the acute management of lactic acidosis and demonstrates its resolution after the discontinuation of linezolid and the use of CVVH. It does mention the patient's condition six days after admission to the ICU and when the patient’s condition was stable, she was transferred to the infectious disease department and antituberculosis drugs started, with the exception of linezolid. This did not result in recurrence of lactic acidosis. The causal relationship between lactic acidosis and linezolid was categorized as ‘probable’ on the Adverse Drug Reac- tion (ADR) Probability Scale. |  |
|  | 8. Is the case described with sufficient details to allow other investigators to replicate the research or to allow practitioners to make inferences related to their own practice? | It reinforces the importance of considering risk factors for linezolid-induced lactic acidosis, such as older age, extended linezolid therapy, and kidney dysfunction.  It suggests that CVVH can be a viable treatment option for managing linezolid-induced lactic acidosis, especially in severe cases. |  |
| Zuber Ahmad et al  (INDIA)  JMS 2023: 26 (2); 38-40.  https://doi.org10.33883/jms.v26i2.1274 | 1. Does the patient represent the whole experience of the investigator (center) or is the selection method unclear to the extent that other patients with similar  presentation may not have been reported? | Patient received treatment for drug resistant extrapulmonary tuberculosis. | Fair Quality |
|  | 2. Was the exposure adequately ascertained? | Yes, the report clearly states the patient received 600mg of linezolid daily for 02 months. This information adequately defines the patient's exposure to linezolid. |  |
|  | 3. Was the outcome adequately ascertained? | The report clearly documents that the patient developed lactic acidosis. Her blood gas analysis showed a pH of 6.98, PaCO2 15 mmHg, PaO2 153 mmHg, , Sodium 141 mmol/L, potassium 4.5 mmol/L, lactate more than 15 mmol/L, and base excess of -26.4 mmol/L. These lab values, along with her symptoms, confirm the outcome of lactic acidosis. |  |
|  | 4. Were other alternative causes that may explain the observation ruled out? | Yes, they made the diagnosis of it as a diagnosis of exclusion whereby ruling out other causes of it. |  |
|  | 5. Was there a challenge and/or rechallenge phenomenon? | No |  |
|  | 6. Was there a dose-response effect? | NO |  |
|  | 7. Was follow-up long enough for outcomes to occur? | Patient died after she developed atrial fibrillation |  |
|  | 8. Is the case described with sufficient details to allow other investigators to replicate the research or to allow practitioners to make inferences related to their own practice? | Yes |  |
| Poobalan Naidoo et al.  (South Africa)  *Wits Journal of Clinical Medicine,* 2023, 5(2) 133–136  http://dx.doi.org/10.18772/26180197.2023.v5n2a8 | 1. Does the patient represent the whole experience of the investigator (center) or is the selection method unclear to the extent that other patients with similar  presentation may not have been reported? | The index case was a 37-year-old female with a back-ground of HIV, on a fixed drug combination antiretroviral treatment consisting of tenofovir, lamivudive and dolutegravir for an unknown duration. She was virologically supressed, with a CD4 count of 619 cells/mm3. Her retroviral disease was complicated with multidrug resistant pulmonary tuberculosis (MDR-TB). | Good quality |
|  | 2. Was the exposure adequately ascertained? | Yes, The exposure to linezolid was adequately ascertained. The case study describes the patient's treatment history, including two separate instances of linezolid exposure: an initial course of 600 mg daily for 2 months and a subsequent course as part of a rescue regimen 1 year later. The exact duration of the second exposure before the onset of symptoms was 5 weeks |  |
|  | 3. Was the outcome adequately ascertained? | The outcome, lactic acidosis, was also adequately ascertained. The patient presented with a lactate level of 12 mmol/L, which peaked at 19 mmol/L. The case report clearly attributes the hyperlactatemia to linezolid after ruling out other potential causes |  |
|  | 4. Were other alternative causes that may explain the  observation ruled out? | The case report states that the patient had no significant renal or liver disease, was not septic, and was well-perfused, leading the authors to conclude that linezolid was the cause of the hyperlactatemia |  |
|  | 5. Was there a challenge and/or rechallenge  phenomenon? | This case does present a rechallenge phenomenon. The patient experienced lactic acidosis upon re-exposure to linezolid after an initial course without adverse events |  |
|  | 6. Was there a dose-response effect? | The case report does not provide information about a dose-response effect. The patient received the same dose of linezolid (600 mg daily) during both exposures |  |
|  | 7. Was follow-up long enough for outcomes to occur? | Unfortunately, the follow-up duration cannot be determined from the information provided. The case report focuses on the acute presentation and management of lactic acidosis, ultimately leading to the patient's death. The report details that the patient went into cardiorespiratory arrest and died despite four hours of continuous veno-venous haemodialysis |  |
|  | 8. Is the case described with sufficient details to allow other investigators to replicate the research or to allow practitioners to make inferences related to their own practice? | The case report provides sufficient detail for other investigators to make inferences related to their practice. It describes the patient's medical history, the timeline of linezolid exposure, the clinical presentation of lactic acidosis, the management strategies employed, and the ultimate outcome. The authors also discuss the rarity of linezolid-induced lactic acidosis, the mechanism of toxicity, and risk factors for its development, providing valuable context for clinicians. |  |
| Shubham Singh et al.  (INDIA)  10.4103/JNMO.JNMO_17_24 | 1. Does the patient represent the whole experience of the investigator (center) or is the selection method unclear to the extent that other patients with similar  presentation may not have been reported? |  | Fair quality |
|  | 2. Was the exposure adequately ascertained? | The report details the 46 years old male patient's exposure to linezolid, including the dosage and timing of administration. The patient received an all-oral longer antitubercular treatment (ATT) regimen that included bedaquiline, levofloxacin, linezolid, clofazimine, and cycloserine. The patient began experiencing symptoms approximately four hours after ingesting the first dose of the ATT regimen. |  |
|  | 3. Was the outcome adequately ascertained? | The outcome, lactic acidosis, was confirmed through arterial blood gas analysis, which revealed a pH of 6.09 and a lactate level of 17.52. |  |
|  | 4. Were other alternative causes that may explain the observation ruled out? | It also states that the patient and family members denied ingestion of any toxic substances or alcohol. The report does not provide details on ruling out other potential causes of lactic acidosis, such as sepsis. |  |
|  | 5. Was there a challenge and/or rechallenge phenomenon? | There is no mention of a challenge or rechallenge phenomenon in this specific case report |  |
|  | 6. Was there a dose-response effect? | Can not be ascertained as patient after receiving first dose of LNZ met with fatal adverse event. |  |
|  | 7. Was follow-up long enough for outcomes to occur? | The patient's condition deteriorated rapidly, and died within 12 hours of symptom onset, making follow-up irrelevant in this case. |  |
|  | 8. Is the case described with sufficient details to allow other investigators to replicate the research or to allow practitioners to make inferences related to their own practice? | The report emphasizes the need to be vigilant for early signs and symptoms of lactic acidosis in patients on linezolid therapy, especially those on a multi-drug regimen. Even a single dose can cause fatal LA. |  |
| Venkat Ramesh et al  (INDIA)  BMJ Case Rep 2024;17:e259335doi:10.1136/bcr-2023- 259335 | 1. Does the patient represent the whole experience of the investigator (center) or is the selection method unclear to the extent that other patients with similar  presentation may not have been reported? |  | Good quality |
|  | 2. Was the exposure adequately ascertained? | The report clearly describes the patient's exposure to linezolid, including the dosage (600 mg daily) and duration (5 months). The patient was on a treatment regimen for disseminated multidrug-resistant tuberculosis, which included bedaquiline, levofloxacin, linezolid, cycloserine, and clofazimine |  |
|  | 3. Was the outcome adequately ascertained? | Yes, the report confirms the outcome, linezolid-induced lactic acidosis (LILA), through clinical findings and laboratory data. Clinically, the patient presented with progressively worsening shortness of breath and vomiting. Laboratory investigations revealed a pH of 7.287, a lactate level of 10.2 mmol/L, and an anion gap of 19 mmol/L4. These findings, alongside the exclusion of other potential causes, support the diagnosis of LILA |  |
|  | 4. Were other alternative causes that may explain the  observation ruled out? | The report describes a systematic process of ruling out other common causes of the patient's symptoms and laboratory abnormalities.  Ruled out Sepsis, Cardiogenic or Obstructive Shock, HAGMA, Metformin Poisoning and Malignancy |  |
|  | 5. Was there a challenge and/or rechallenge  phenomenon? | There is no mention of a challenge or rechallenge phenomenon in this case report |  |
|  | 6. Was there a dose-response effect? | The dose-response effect of linezolid on lactic acidosis is not explored in this case report |  |
|  | 7. Was follow-up long enough for outcomes to occur? | The patient's condition improved after discontinuing linezolid and receiving supportive care. The report mentions the patient was doing well on follow-up treatment and completed 6 months of bedaquiline. However, the exact duration of follow-up is not specified |  |
|  | 8. Is the case described with sufficient details to allowother investigators to replicate the research or to allow practitioners to make inferences related to their own practice? | The case study provides a detailed account of the patient's medical history, clinical presentation, laboratory findings, treatment, and outcome. This information allows other investigators to make inferences related to their practice. However, the report does not mention any specific details that would allow for direct replication of the research. |  |
